# Correcting Covid-19 PCR Prevalence for False Positives in the Presence of Vaccination Immunity

**DOI:** 10.1101/2021.04.06.21255029

**Authors:** Michael Halem

**Affiliations:** BeCare Link LLC

**Keywords:** Epidemiology, PCR, False Positive Rate, FPR, Vaccination, Model, Prevalence, Decay, Exponential

## Abstract

Since the first analysis was published on 7 April 2021 the PCR test positivity rate has dropped significantly below the then estimated false positive rate (FPR) of 1.16% using the exponential decay to FPR model. Therefore, the estimate has been rejected and a new model was developed.

Using the ONS infection survey’s assumption (PCR FPR rate below 0.1%) the new model splits the test time series data into two periods based on a change in transmissibility that coincides with the reopening of England schools on 8 March. The new model provides for two base levels of exponential decay (for each period’s transmissibility) combined with a single decay rate increase dependent on vaccination.

Because the FPR is relatively insignificant compared to current PCR test positives, it cannot be statistically separated using currently available England epidemic time series data by the non-linear least squares estimation technique. Therefore, the FPR factor is temporarily dropped in the least squares regression.

The new model is stable in that it reasonably predicts through the most current available data (25 April) the future test prevalence using parameters estimated with 29 March data. Thus far, the estimate parameters remain within their original confidence intervals as successive days are added to the time series. Of potential usefulness is the current estimate for change in decay rate per mean vaccination rate, currently estimated at approximately 10.7% (CI: 8.8% - 12.6%). The estimate should be used with caution as other unforeseen factors could cause the model to misestimate.

## Introduction

The end of an epidemic, due to either constant large transmission reduction (i.e. *R* ≪ 1) or by a new removal of a large fraction of the susceptible population due to vaccination, would be expected to be an exponential decay curve under SIR assumptions. Test positivity time series would reasonably be expected to resemble an exponential decay, with a floor at the test false positive rate. However, when the test population positive prevalence greatly exceeds the test false positive rate, the FPR floor cannot be discerned by estimation techniques based on statistical time series. Where FPR ≪ Prevalence due to other evidence, FPR can be ignored in exponential regression based models until the FPR becomes a statistically significant portion of the test population prevalence.

An earlier version of this paper [1] used an FPR floor regression model which captured a statistical artifact, a false FPR floor. This right side tail of the 29 March time series data coincided with the return of English school pupils to the classroom for in-person learning on 8 March 2021 [2]. As can be clearly seen in the results section the positivity fell significantly below the FPR floor. Therefore, the floor must be clearly below the 1.16% found by the 7 April paper’s model, or the 1.3% found by a revised FPR model which both corrected the test time series to un-bias for case de-duplication and LFD positives added to the PCR test sample. Note: the revised model also an erroneous vaccinations with time acceleration factor^†^. As such, the FPR floor model’s results (and estimates) are rejected for this time period.

Therefore, a new model was developed. The regression is divided into two periods, before and after the return to school. It is shown that the fit both improves and is stable through time, thereby explaining with only four regression factors: initial prevalence, initial decay rate at the start of the first period, initial decay rate at the start of the second period, and the change of the decay rate as a percentage of the mean vaccination immunity over the time period. To make the transmissibility consistent, the initial data period was started on 6 January (instead of 1 January) because 6 January is the date that the stay at home order came into force for England [3].

This paper and the accompanying code provides a technique for estimating these regression factors. It also provides the original method (as corrected^†^, using optionally one or two periods) for estimating the FPR for future use at such time as the test population prevalence approaches the FPR.

## Methods: Data

For the analysis, it is necessary to have a relatively clean data stream with biases removed. For most public reporting, the PCR and LFD (lateral flow device) test results are either combined to give case counts; or if kept separate, the PCR test results may be biased using reported LFD positives without adjustment for the total LFD tests conducted in the denominator. Further, most Public Health England (PHE) test reporting is “de-duplicated” so that once an individual is tested positive, subsequent positive tests are not recorded in the data stream.

The data is downloaded from PHE as seven day rolling total PCR, LFD, and non-rolling daily vaccination data as csv files [4, 5]. PHE fieldnames are in **bold** below.

Fortunately, PHE has provided separate data fields for PCR tests and LFD tests, including for LFD tests that have been confirmed by PCR. This data is also de-duplicated, such de-duplicated tests being termed as “cases” by PHE. To provide an estimate of the PCR test positivity, a comparable test numerator and denominator must be computed which includes duplicate (repeat) tests on an individual. It is assumed that the positivity ratio of all tests is approximately equal to the positivity ratio of the cases. To avoid LFD positivity bias of the PCR stream

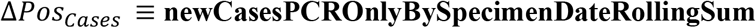

is used as the numerator. Note that all PHE provided rolling sums are divided by 7 to provide a 7 day trailing moving average. The above “cases” field is de-duplicated, thus a de-duplicated denominator must be computed.

Because there is no method of estimating the total positive tests from the numerator, instead, a de-duplicated daily moving average of total PCR test for the denominator (of the positivity percentage) is estimated from other available time series; thus providing a ratio of de-duplicated positive tests to de-duplicated total tests. (Ideal data would provide both these time series for all PCR tests, not de-duplicated, and not biased for positive LFD tests. The research of the author has not found this data available.)

The estimated total PCR daily tests is estimated from:

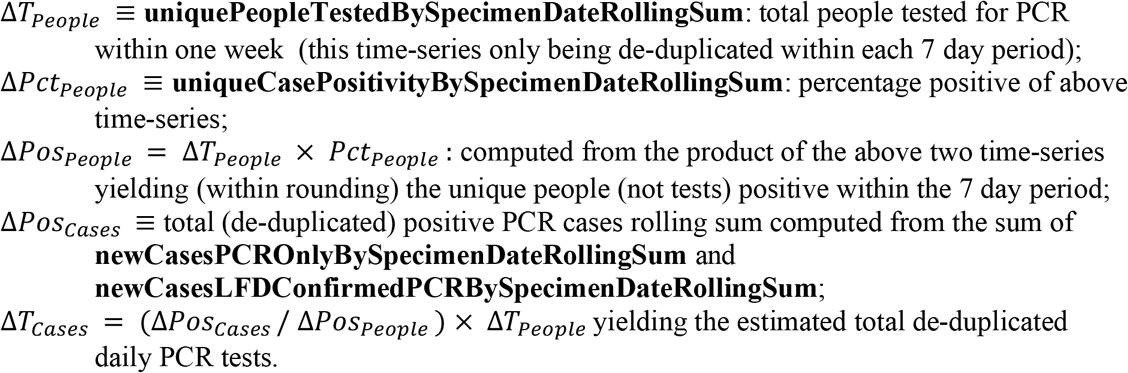

The total PCR positivity percentage estimate, unbiased for LFD test is thus:

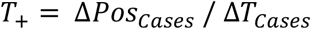

Note: the denominator of total PCR tests can be adjusted by subtracting out the total (positive) LFD tests that have been submitted to PCR for testing. However, experimenting with the data shows that adding this denominator correction has no effect on the accuracy of the predictions as the difference of the mean is only 0.03% and the standard deviation is also 0.03%, such that when plotted on a graph the data points all appear to exactly overlap. (The attached code has this adjustment as an option.) Because the PHE policy has changed in early April, to after the policy change, submitting positive LFD tests for PCR verification as part of wider LFD screening, this correction was not used in the subsequent analysis to remain consistent over the entire January through April time series.

The test positivity approximation was double-checked by downloading the weekly ONS Pillar 2 Test Conducted Spreadsheet (i.e. tests_conducted_2021_04_22v2.ods, Table 1)[6], showing the total PCR test taken and positive test results for the week, and then unbiasing the Pillar 2 positive tests for LFD positives that have been confirmed by PCR in the numerator (from the daily data downloaded previously above), and then unbiasing the denominator by total LFD test submitted to PCR, to yield an unbiased weekly estimate of Pillar 2 tests and positives. Pillar 1 testing was unbiased using the weekly Test and Trace spreadsheets (i.e. NHS_T_T_data_tables_Output_Week_46.ods, Table 1)[7], to yield a combined Pillar 1 and Pillar 2 unbiased weekly testing numerator and denominator. The double-check was done with the then available weekly data, from 6 January thru 14 April, and visually the weekly data points line up on top of the daily moving average time series used for the regressions in this study. Because the two spreadsheets only provide weekly data, they yield only 15 weekly data points which are insufficient for a regression with 4 coefficients to be determined and cannot be used directly, but only as a double-check.

Finally, vaccination data is downloaded:

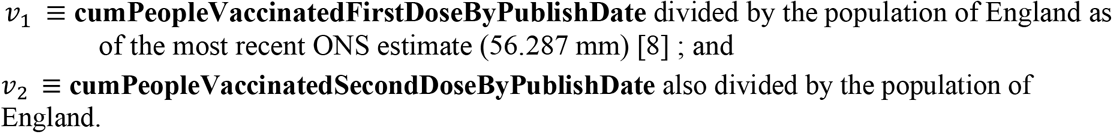

From these the effective vaccination coverage is computed on each date assuming 80% effective immunity from the first dose [9] and an additional 20% effective immunity from the second dose; with the immunity level based upon the vaccinations published 7 days prior, i.e.

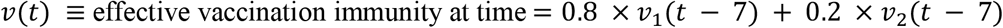

## Methods: Modelling

An inspection of the current test data will show that there is a decline similar to exponential decay as a large portion of the English population became vaccinated. The precise decay dynamics is not needed as the exponential model is sufficiently parsimonious and consistent with solutions to SIR family of models when the susceptible population has been depleted sufficiently for the effective reproduction number R to be significantly below 1 over short time periods. Generally, a population’s currently infected prevalence exponential decay (or growth) can written as:

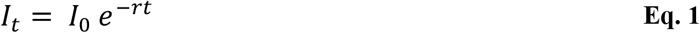

where *I*_*t*_ is the percentage prevalence at time *t* (i.e. infectious, or previously infected depending on the authorities definition and testing strategy), *I*_1_ is the initial prevalence at the start of the measurement period, *t* is the days from the start, and *r* is the daily decay rate (which for brevity is a positive number in this paper). The exponential solution follows directly from the differential equation for the change of infection with respect to time in the classic SIR model and similar models [10]

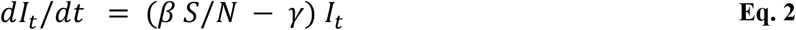

where *I* is the time varying infection, *t* is time (by convention in days), *β* is the transmissibility (assumed constant), *S*/*N* is the percentage of population that is susceptible, and *γ* is the recovery rate from the infected group to the recovered group. Using the exponential distribution, *γ* = 1/*τ*, where τ is the mean time of infection: for COVID-19 somewhere between 5 and 10 days. It can be seen by substitution that Eq. 1, the exponential increase or decay, is the solution to Eq. 2 when all parameters in the parenthesis are constant. Further it can be seen that the rate of exponential decay −*r* = *β S*/*N* − *γ*. From inspection it is obvious that the decay rate r is a linear function of the susceptible percentage of the total population *S*/*N* such that over shorter time periods, and with assuming transmissibility *β* is constant and where new natural infections are small relative to vaccinations, the decay rate is a linear function of the vaccinations that have removed susceptible people from the population.

This allows us to write a general revised version of *r* for a regression model and substitute it into the infection decline equation Eq. 1 :

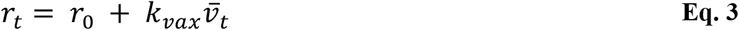

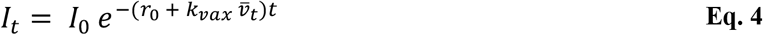

where *r*_0_ is the initial decay rate at the start of the period (defined as *t* = 0), inclusive of the effect of any vaccinations, 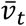 is the arithmetic mean percentage with vaccination immunity for the time interval 0 to *t*, and *k*_*vax*_ is a regression solved constant showing the excess decay per mean effective vaccination over the period.

For the situation where the transmissibility is relatively constant but then changes at a known time *t*_1_ to a different level, more consistent results can be obtained by dividing the epidemic curve *T*_+_ into two piecewise continuous segments with a single vaccination *k*_*vax*_ constant. This is done by starting the second segment at the end mark for the infection in the first segment, and adjusting the vaccination rate to avoid double counting of vaccinations from the first period in the second period. Using this convention, both *r*_0_ and *r*_1_ are the infection decay rate as if there was no vaccinations. *I*_*t*1:*t*0_ in Eq. 7, the infections at the end of the first period is the transition point, the start for the second time period (Eq. 6).

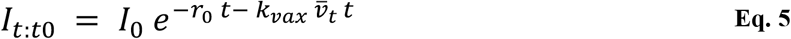

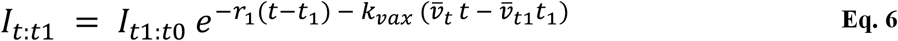

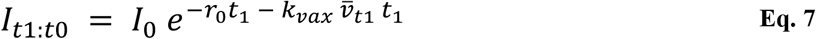

With algebra and the use of minimization and maximization functions min() and max(), the combined 2 period infection rate can be reduced to a single equation for regression so that the computation is correct for both the initial period (Eq. 5) and the second period (Eq. 6):

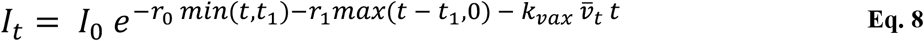

The algebra for computing test positivity from population prevalence is well known. For a complete derivation please see [11] mathematics appendix, where Equation 30A is presented here as Eq. 9 under the simplification that *N* = 1 so that all units are in percentage of the population. Using the definition of specificity as 1 − *f*, where *f* is the false positive rate (FPR) then

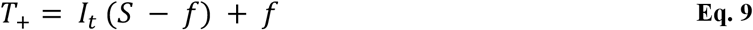

where *I* is the population prevalence, *T*_+_ is the test positivity, and *S* the test sensitivity. With no reference standard, the sensitivity is not estimated and is set arbitrarily to 1 (i.e. perfect):

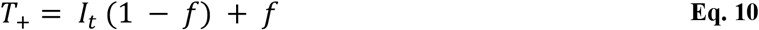

An overall model, suitable to solve with empirical test data using R’s non-linear least squares error minimization techniques^‡^ [12, 13, 14] is created by substituting *I*_*t*_ into equations Eq. 10 from either Eq. 4 (single period model) or Eq. 8 (two period model):

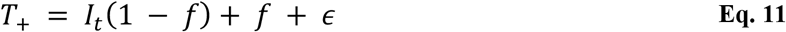

where *ϵ* is the residual error that the non-linear least squares model is minimizing. In the case where the false positive rate *f* ≪ *I*_t_ the *f* term is dropped (i.e. assumed to be zero). To equally weight the percentage error as the decay progresses with test time within the least squares minimization algorithm, the system is solved by taking the log of both sides, thus preventing excessive minimization on the larger absolute errors at the beginning of the regression time frame:

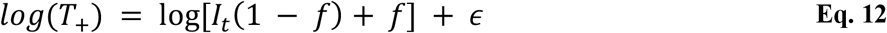

Note that in the case where the *f* term is dropped, i.e. where the FPR is not being estimated, the right side can be solved using an ordinary least squares linear regression to minimize the error in the *log*(*T*_+_) estimate (as the log and the exponent functions cancel leaving a linear equation in the right hand side in *r*_0_, *r*_1_, *k*_*vax*_ and with *log*(*I*_0_) being the regression constant. Also note that *k*_*vax*_ is computed on the product 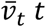, the later (product) being a constant at every sampled time point.

## Results

The result is summarized below:

**Table 1.**
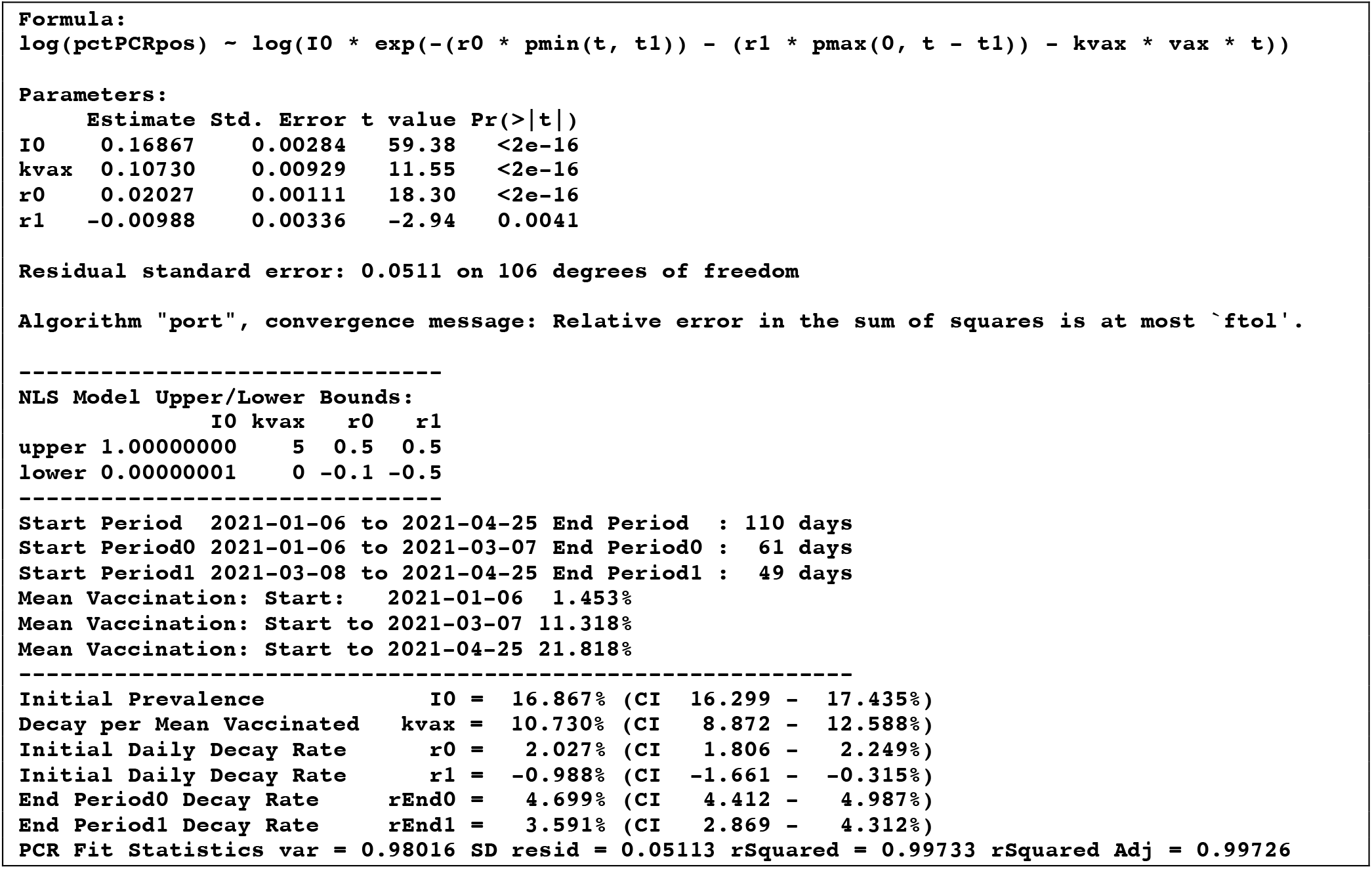
Model Fit Result.

The estimates and their confidence intervals (+/- 2 standard errors) have been extracted to 6 decimal places of precision, with an ad hoc *R*^2^ (percent of the variance of the independent variable explained) so as to be comparable to an ordinary least squares linear regression.

As this model fit did not estimate a false positive rate, it is assumed as zero such that the test prevalence *I* is the same as the test positive rate *T*_+_. The regression estimates l*og*(*T*_+_) via the R predict.nls() function (part of the base R stats package). *T*_+_ is estimated as 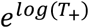 producing the test population prevalence as *I* = *T*_+_ by rearranging Eq. 10 to Eq. 13 and assuming *f* is zero:

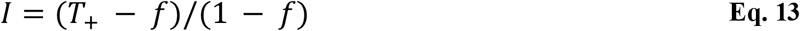

Confidence intervals were obtained when directly estimated by the R nlsLM() function, by doubling the nlsLM() summary standard errors. As the data is entirely based on 7 day moving averages^§^, daily estimates of fit data points would have approximately 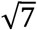 times wider confidence intervals. However, the regression fit coefficients represent the fit to 7 day moving averages and are therefore consistent estimates on that basis. The ending decay rates for each period (*r*_*end*0_ and *r*_*end*1_) were estimated assuming that variables were random, normally distributed, and uncorrelated using the simplifying formula for the variance of the addition of two such random variables, i.e. 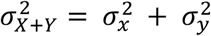.

In the case of the end of period population prevalence estimate *I*_*end*_, the regressions estimate of *log*(*T*_+_) has a residual standard error, in this case 0.0511. The CI for *T*_+_ is thus 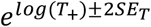. The confidence interval was calculated by substituting into Eq. 13 the respective worst case standard errors for *T*_+_ and *f* (i.e. flip the sign for the plus or minus):

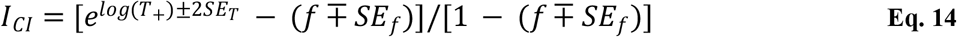

Of note, because the PCR sensitivity is set to 1 and because the threshold for infectiousness is around Cycle Threshold (Ct) 32 while ONS has reported that its testing program using Ct’s that exceed 37, the testing may well find positive samples that are not infectious, i.e. viral fragments from prior infections or even viral fragments or single virions filtered from other nearby infectious individuals but not infecting the tested subject. This high sensitivity would thus give the test results a tendency to include past infections as a kind of extended rolling sum window. While these are not false positives from a testing standpoint, it would have a tendency to decrease the decay rate as the non-infectious particles are delayed in clearance from the population over time.

As a check against the stability of the model, it was also run for the period ending 29 March -- the cutoff date for the prior version of this paper, to predict against the most current data (through 25 April) as of the time of this writing (30 April):

**Table 2.**
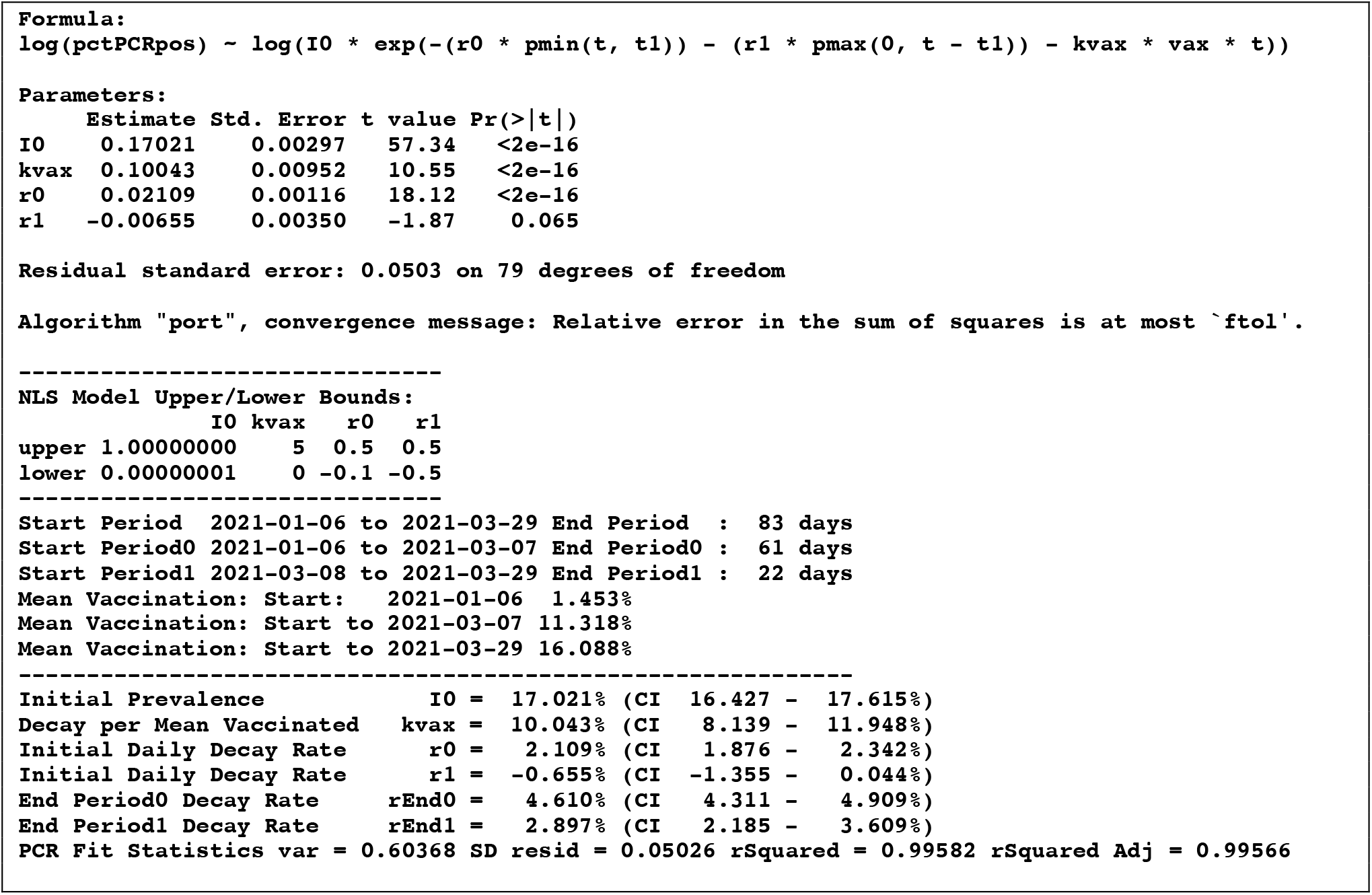
Model Fit Result.

From an observation of the parameter estimations, it can be seen that the 25 April estimates are all within the confidence intervals of the 29 March estimates. This can be seen graphically by comparing the two estimates shown in Figure 1:

**Figure 1.**
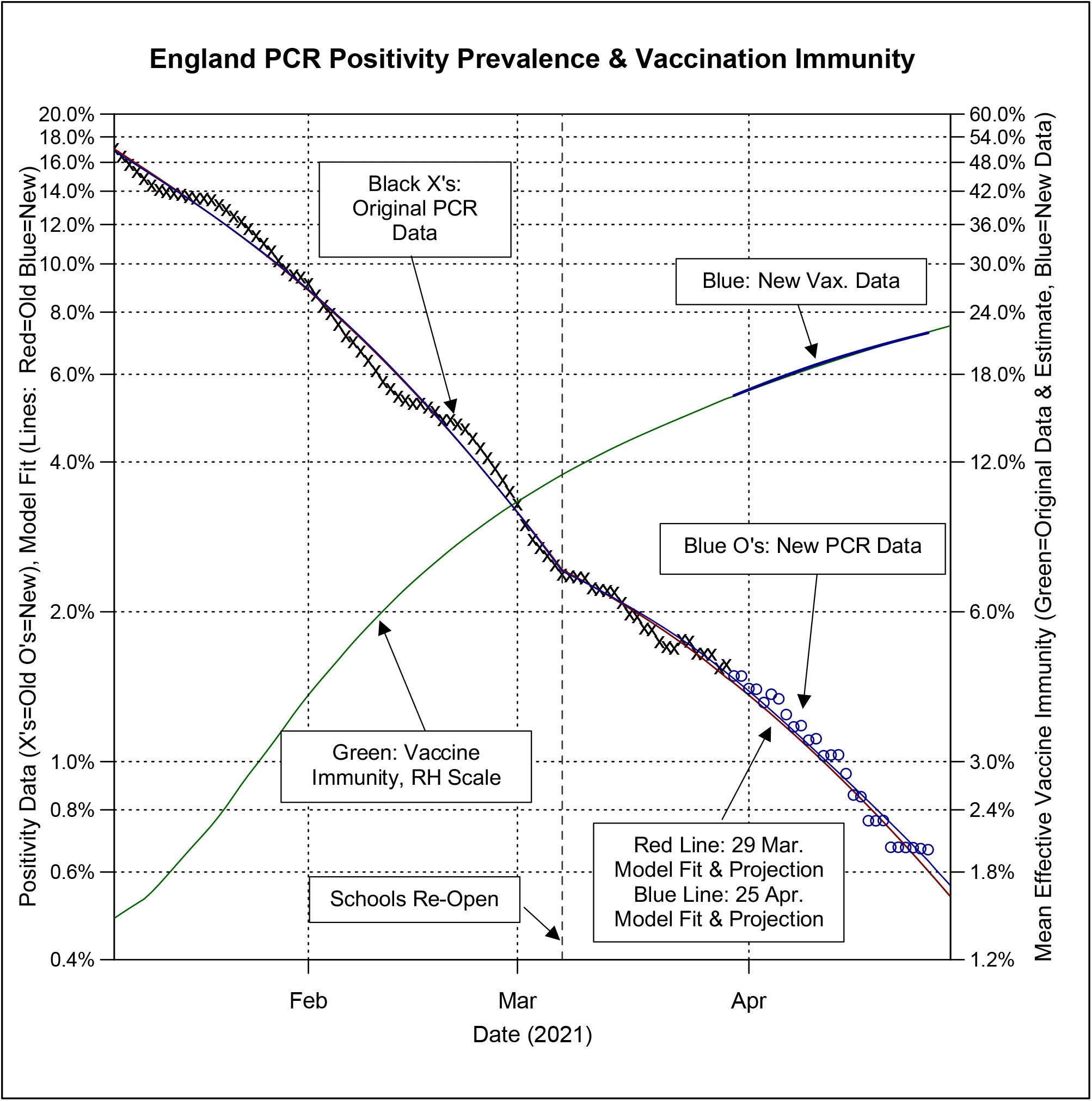
Revised 2 Period Model Fits & Projection. The original model, shown as a red line through the data points was fit only to the original data (the Black X’s). However, for the 27 day period where new data has become available, the original model fit still closely matches the new data points. Of note is that a new model fit based on the entire data set, shown as the blue line, almost exactly overlays the original red line fit to make the line appear black to by around mid-March, indicating very little model drift.

The prior version of this paper included an analysis which explicitly included a false positive rate parameter in the non-linear least squares regression for a single period. The subsequent flattening of the test positivity time series after this reopening caused the model to inaccurately find a floor which the model interprets as a false positive rate (FPR) for the PCR test of over 1%. As can be seen in Figure 2 clearly this estimate is too high as the test positivity subsequently descended through the FPR floor.

**Figure 2.**
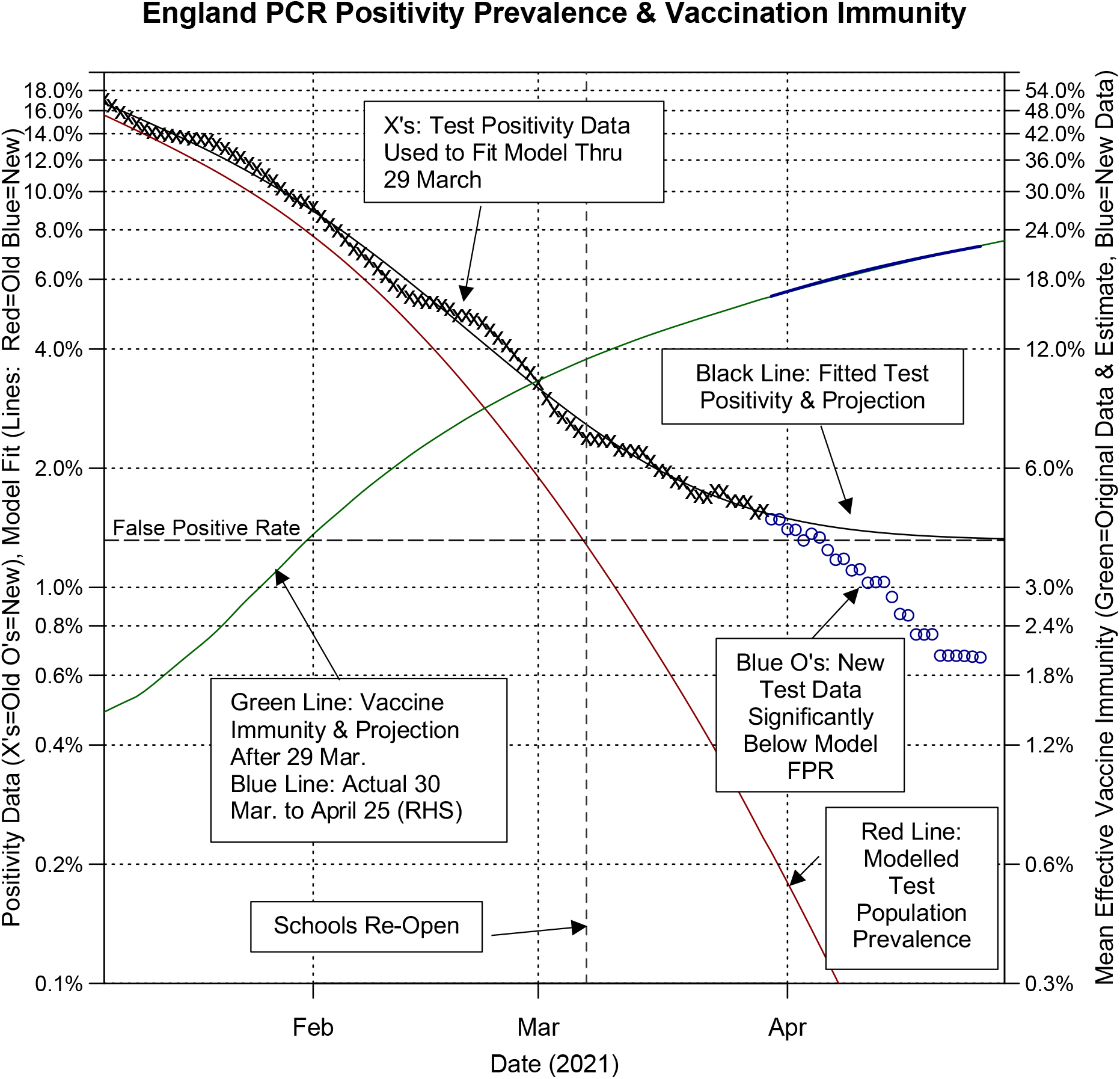
Single Period Model Incorporating FPR Parameter. Single Period Model Incorporating FPR Parameter This model is similar to the one used in the 7 April This model is similar to the one used in the 7 April version of this paper. It can clearly be seen that subsequent data points (the blue O’s) have fallen significantly below the false positive rate, which violates the initial model assumptions. Therefore, the actual PCR test FPR rate must be significantly below the >1% threshold found by the model. Of note is that 1) the data de-biasing has been revised so that the unbiased PCR test positivity is somewhat higher than previously; and 2) the current model no longer multiplies the vaccination by the square of time in the decay rate as can be seen by comparing Eq. 3 to the 7 April version of this paper. The results are similar but not identical.

## Discussion and Conclusion

A technique is presented to extract the effect of vaccination on the decay rate of an epidemic. An additional technique is made available whereby the test false positive rate can be included. However, until such time as the test population prevalence falls to a rate that is within a reasonable factor, say less than two times the test false positive rate, the model may falsely capture a false positive floor that is due to other unrelated factors.

Most recently ONS has published [15] an update on their estimate of PCR specificity (i.e. FPR) where they believe the FPR must be 0.08% or below. They further cite a paper by an academic consortium (Walker et al.) [16] which specifically diagrams the relatively high cycle thresholds above 35 for newer single gene variants. By extrapolating their Supplementary Figures, it is possible to show that at these high cycle thresholds (Ct’s) single copies per ml can trigger a positive test. This suggests that the positivity window is rather wide compared to the expected period of infectiousness, meaning that test positivity data will lag the termination of actual infections by a window. This may possibly effect the exponential decay time series, to either broaden the test reported peaks and troughs, or alternatively to create a complication in the exponential decay if the mean removal of post infectious particles from test subjects itself has an different exponential decay. This complication is beyond the scope of these relatively simple regression models.

The most remaining useful item from this analysis is the estimate of the increase in the decay rate from vaccination acquired immunity — currently estimated at around 10.7% with a confidence interval between 8.9% and 12.6%. While this appears relatively stable, it is possible that the current model has also latched onto a new, different data artifact, so this estimate should be used with caution until confirmed independently. Its units are change of rate of decline of infection per mean vaccine immunity over the entire period. Mean vaccine immunity can be increased by increasing vaccination, and it indicates a limit of approximately 10 to 11% in the rate of decline that can be obtained by vaccinating 100% of the population. This may be related to the average rate of infection clearance in an infected individual, and it may be also limited by the rate of transition from positive to negative of a previously infected individual who is no longer infected, but carries non-infectious single copy virus remnants detected at Ct>37 in their collected sample for an unknown additional period of time.

It should be further noted that there is also, due to taking the derivative of Eq. 4 or Eq. 8 with respect to time, that the chain rule of differentiation provides a temporary increase in decline when 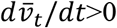, i.e. when the mean vaccination rate increases. As this mean is the average of the last day’s vaccination and all previous days vaccinations (which are past and hence fixed), this will have a tendency to temporarily increase the rate of decline when significant daily vaccinations occur. Otherwise, 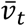 will only slowly increase as over time as the mean vaccination percentage approaches the final vaccination percentage^**^ This implies a small boost in the rate of decline during periods of heavy relative vaccination, with the boost slowly decaying once the vaccination campaign has finished at whatever the final uptake level the population will accept.

### Limitations

This paper is not peer reviewed. While the author exercised reasonable care in presenting the results, hidden mistakes may be contained therein. The statistical techniques used within this paper may not be statistically robust.

This model is a simplification based on the ending state of an epidemic under a SIR or SEIR framework where the decay becomes largely exponential. At some point, the total immunity and current transmissibility can be corroborated against the rate of decline which is an aggregation of these other variables.

While the Public Health England data are relatively clean, it is an amalgamation of multiple PCR testing sites, each which may change its test parameters at any time, resulting in a different false positive rate. The technique performs best when the testing parameters and the tests used are consistent and homogeneous. For example, some (non-England) public health authorities may mix lateral flow test results with PCR test results; or may bias test results by screening first with LFD, and then dropping negative LFD results from the reported time series. Such mixed, changed, or biased data makes the ability of the technique to estimate parameters and to fit the data less reliable on such data.

A constant transmissibility *β* (i.e. social distancing) is assumed except for a single change in the two period model. The change in naturally acquired immunity by infection over each period is assumed to be relatively insignificant. Note that additional periods can be added if additional significant changes in transmissibility occur, e.g. due to reopening.

The technique works during an epidemic curve period when the total infections are falling rapidly in a consistent manner to within a small multiple of the false positive rate, so that the false positive rate floor can be detected. During other periods, the false positive floor may not be discernible to the model, as the statistical sampling and testing random variations and other effects in the population (i.e. changes in transmissibility) may mask a relatively small underlying false positive rate within the data noise.

The reported validation of many COVID-19 PCR tests indicate that the specificity rate is 100% (i.e. false positive rate is zero). [15, 17, 18]. Of note, is that the UK Government Office of Statistics previously stated in April 2020 that the operational false positive rate was unknown [19] and that an April 2020 review [20] of COVID-19 PCR false positives published test results found a wide range (0.3% to 6.3%) of false positive rates for both COVID-19 PCR and other comparable non-COVID-19 PCR tests.

## Supporting information

Code/Data for 2021-04-30 version

## Data Availability

All code and data is on medRxiv under the supplementary-material link as a zip archive. See: https://doi.org/10.1101/2021.04.06.21255029

## Footnotes

† The original model used 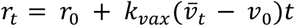 with *t* representing a hypothesized acceleration with time factor. The corrected model uses 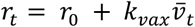, This and the revised data de-biasing provisions did not appreciably change the estimate for FPR based on the original data, going from 1.16% to a revised 1.32%.

‡ Note that the nlsLM() [14] function is more stable, i.e. converges to a solution more reliably, than the base R nls() function.

§ 7 day moving averages are used to smooth out “weekend” effects that otherwise create systematic noise.

** This is a consequence of 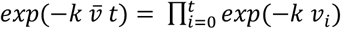, i.e. the total exponential decline versus time due to vaccination is the product of the daily percentage decline due to the identity *exp*(∑ *x*_*i*_) = ∏ *exp*(*x*_*i*_).

## Notes

### Competing Interest Statement

The authors have declared no competing interest.

### Funding Statement

No external funding support. The author did this study independently without funding.

### Author Declarations

IRB - Not applicable.

### Summary of Updates

1) Since the first analysis was published on 7 April 2021 the PCR test positivity rate has dropped significantly below the then estimated false positive rate (FPR. Therefore, the estimate has been rejected and a new model was developed. 2)The new model splits the test time series data into two periods based on a change in transmissibility that coincides with the reopening of England schools. The new model provides for two base levels of exponential decay (for each period's transmissibility) combined with a single decay rate increase dependent on vaccination. 3)Because the FPR is relatively insignificant compared to current PCR test positives the FPR factor is temporarily dropped in the least squares regression. 4) New code provided for all above.

