## Supplementary material for "Correcting Covid-19 PCR Prevalence for False Positives in the Presence of Vaccination Immunity": Code/Data for 2021-04-30 version: correct4falsePos-v2021-04-30.r

```
## Correcting For False Positives Paper R Code
## Paper Title: Correcting COVID-19 PCR Prevalence for False Positives in the Presence of Vaccination Immunity
## version 2021-04-07: For April 7th Paper pushed to medRxiv 
## Version 2021-04-30: For April 30th Paper pushed to medRxiv
## Paper available on MedRxiv https://doi.org/10.1101/2021.04.06.21255029
## Code and Paper Author: Michael Halem 
## Code (c) Michael Halem 2021
## This code may be freely used or reused as long as it is attributed to the author.
####################################################################################
## Startup R and source this file, i.e. > source('correct4FalsePos.r')
## Then
## > fetchEngland()            ## will fetch the data to your download directory.
## Note: URLs are within the fetchEngland() header comment below if the data must be manually downloaded (from PHE) using a browser.
## > doFigure1AndTables2and1() ## Draws figure 1 and prints to console Table 2 and Table 1
## > doFigure2()               ## Draws figure 2
## Other functions:
## getEnglandCorrected()       ## Flexible main analysis routine and graphing routine: many options documented with function below
## getVaxEngland()             ## Read the downloaded England Vaccination Data
## Questions/Requests:

####################################################################################
## Utilities, constants, and other one-time setup
library('minpack.lm')          ## If you don't have this on your system, install from CRAN
dateRefPosix = as.POSIXct('2019-12-31 OO:OO', tz='EST')
DateRefDate = as.Date('2019-12-31', tz='EST')
options(width=266, digits=5)
options(stringsAsFactors=F)
printf<-function(...) cat(sprintf(... , sep=''))
#####################################################################################

## The following functions will draw figure1, print tables 2 and 1 in that order
## This acts serves as brief tutorial on using the analysis function
## Note that 3 newer data points (26-28 April as blue circles and lines) when the PHE API provides them 
## if you run fetchEngland() instead of using the accompanying data files downloaded on 30 April 

doFigure1AndTables2and1<-function()
{
    getEnglandCorrected(nPoints=83, nExtendPlot=30, bDoPlot=T, bFindFPR=F, bOnePeriodModel=F, startDate="2021-01-06", bRetData=F)  
    newFit = getEnglandCorrected(nPoints=110, nExtendPlot=30, bDoPlot=F, bFindFPR=F, bOnePeriodModel=F, startDate="2021-01-06", bRetData=T)  
    with(newFit, points(fit~date, type='l',col='darkblue'))     
}

## This function will draw figure 2 from the paper.
doFigure2<-function()
{
    getEnglandCorrected(nPoints=83, nExtendPlot=30, bDoPlot=T, bFindFPR=T, bOnePeriodModel=T, startDate="2021-01-06", bRetData=F)      
}
    

#####################################################################################
## Fetch today's data from PHE website to disk.  Note, only run a few times per day to avoid being throttled by PHE.
## If you cannot get this to work on your machine (missing curl command or doesn't understand unix style paths),
## you can instead save the file directly by
## going to this link and chosing "save as" englandData.csv as your file in your Downloads folder.
## Note: If the fields have chnaged in szFetch1-5 below update the URL to fetch those fields.
## https://api.coronavirus.data.gov.uk/v2/data?areaType=nation&areaCode=E92000001&metric=newCasesPCROnlyBySpecimenDateRollingSum&metric=newCasesLFDConfirmedPCRBySpecimenDateRollingSum&metric=newCasesLFDOnlyBySpecimenDateRollingSum&metric=newPCRTestsByPublishDateRollingSum&format=csv
## and for the vaccine data, save as englandVaccine.csv from the following URL:
## https://api.coronavirus.data.gov.uk/v2/data?areaType=nation&areaCode=E92000001&metric=cumPeopleVaccinatedFirstDoseByPublishDate&metric=cumPeopleVaccinatedSecondDoseByPublishDate&format=csv
## Note: The api only allows fetching of 5 fields at a time.

fetchEngland<-function(szPath="~/Downloads")
{
    szFetchPre = 'https://api.coronavirus.data.gov.uk/v2/data?areaType=nation&areaCode=E92000001&'
    szFetch1 = 'metric=newCasesPCROnlyBySpecimenDateRollingSum&'
    szFetch2 = 'metric=newPCRTestsByPublishDateRollingSum&'
    szFetch3 = 'metric=newCasesLFDConfirmedPCRBySpecimenDateRollingSum&'    
    szFetch4 = 'metric=uniqueCasePositivityBySpecimenDateRollingSum&' ##sadly, only to 2 digits
    szFetch5 = 'metric=uniquePeopleTestedBySpecimenDateRollingSum&'
    szFetchPost = 'format=csv'
    szFileName = 'englandData.csv'
    szFileName = sprintf("%s/%s", szPath, szFileName)
    szFetch = sprintf("%s%s%s%s%s%s%s", szFetchPre, szFetch1, szFetch2, szFetch3, szFetch4, szFetch5, szFetchPost)
    download.file(url=szFetch, destfile=szFileName, method='auto')
    printf("... Downloaded to %s\n", szFileName)

    ##Because we now need six non-vaccine fields, have added LFD Only to this file
    szFetchPre = 'https://api.coronavirus.data.gov.uk/v2/data?areaType=nation&areaCode=E92000001&'
    szFetch1 = 'metric=cumPeopleVaccinatedFirstDoseByPublishDate&'
    szFetch2 = 'metric=cumPeopleVaccinatedSecondDoseByPublishDate&'
    szFetch3 = 'metric=newCasesLFDOnlyBySpecimenDateRollingSum&'    
    szFetchPost = 'format=csv'
    szFileName = 'englandVaccineAndMisc.csv'
    szFileName = sprintf("%s/%s", szPath, szFileName)
    szFetch = sprintf("%s%s%s%s%s", szFetchPre, szFetch1, szFetch2, szFetch3, szFetchPost)
    download.file(url=szFetch, destfile=szFileName, method='auto')    
    printf("... Downloaded to %s\n", szFileName)

    ##Old fields fetched in previous versions
    ##szFetch1 = 'metric=newCasesPCROnlyBySpecimenDateRollingSum&'
    ##szFetch2 = 'metric=newCasesLFDConfirmedPCRBySpecimenDateRollingSum&'
    ##szFetch3 = 'metric=newCasesLFDOnlyBySpecimenDateRollingSum&'    
    ##szFetch3 = 'metric=newPillarOneTestsByPublishDate&'  ##Only 5 can be fetched at a time
}


####################################################################################
## Read from file and return the England data for the analysis into a data.frame

getEnglandData<-function(startDate = "2021-02-22", fname="~/Downloads/englandData.csv",
                         fname2 = "~/Downloads/englandVaccineAndMisc.csv",
                         bDebug=FALSE,
                         bCorLFDDenom=T) ##Correct for LFD tests in the denominator
{
    dat = read.csv(fname, stringsAsFactors=FALSE)
    datX = read.csv(fname2, stringsAsFactors=FALSE)
    dat$date = as.Date(dat$date)
    datX$date = as.Date(datX$date)
    dat$newCasesLFDOnlyBySpecimenDateRollingSum = rep(NA, dim(dat)[1])
    imatch = match(dat$date, datX$date)
    for (i in 1:dim(dat)[1]) ##fill in the overflow from datX
    {
        if (!is.na(imatch[i]))
            dat$newCasesLFDOnlyBySpecimenDateRollingSum[i] = datX$newCasesLFDOnlyBySpecimenDateRollingSum[imatch[i]]
    }
           
    dat = dat[order(dat$date),]
    
    ## the fetch should be only for England, so that this field match is superfluous
    dat = dat[,-match(c('areaType','areaCode','areaName'),colnames(dat))]
##    dat$PCRpos = dat$newCasesPCROnlyBySpecimenDateRollingSum/7 ## + dat$newCasesLFDConfirmedPCRBySpecimenDateRollingSum/7
    dat$dC.PCR = dat$newCasesPCROnlyBySpecimenDateRollingSum/7 + dat$newCasesLFDConfirmedPCRBySpecimenDateRollingSum/7
    dat$dT.PCRP =  dat$uniquePeopleTestedBySpecimenDateRollingSum/7
    dat$dC.PCRP = 0.01*dat$uniqueCasePositivityBySpecimenDateRollingSum*dat$dT.PCRP
    dat$dT.PCR = dat$newPCRTestsByPublishDateRollingSum/7
    dat$dT.PCR = c(dat$dT.PCR[-(1:2)], NA, NA) ##adjust for PCR being two days behind
    dat$dT.NoRep = dat$dC.PCR/dat$dC.PCRP * dat$dT.PCRP
    dat$dC.LFD = dat$newCasesLFDConfirmedPCRBySpecimenDateRollingSum/7 + dat$newCasesLFDOnlyBySpecimenDateRollingSum/7
    ##EXPERIMENT... dat$PCRtstPubDate = dat$newPCRTestsByPublishDateRollingSum/7
    dat$PCRtst = dat$uniquePeopleTestedBySpecimenDateRollingSum/7 ##this is a PCR data field.  Because all positive tests are de-duplicated per person, this may be a better denominator base

    if (FALSE)
        {
            dat$pillar1 = movAvgTrail(dat$newPillarOneTestsByPublishDate)
            dat$pctPil1 = dat$pillar1/(dat$newPCRTestsByPublishDateRollingSum/7)
        }
    else
        dat$pctPil1 = rep(NA, dim(dat)[1]) ##this is because only 5 fields can be fetched at once
    ##Published date represents speciments collected about 2 days previously.
    ##Note this 2 days is the best fit of PHE PCR _people_ by specimen data vs the above PCR tests (not people) by publish date
    ##Experiment dat$PCRtst = c(dat$PCRtstPubDate[-(1:2)], NA, NA) ##converting published date to specimen date
    dat$pctPil1 = c(dat$pctPil1[-(1:2)], NA, NA)
    
    ##dat$PCRtst = dat$PCRtstPubDate
    if (!is.null(startDate))
    {
        startDate = as.Date(startDate)
        dat = dat[dat$date>=startDate,]
    }
    ##Now, adjust the denominator to remove all _positive_ LFD tests (assumed passed to PCR testing)
    ##dat$PCRtst = dat$PCRtst - (dat$newCasesLFDConfirmedPCRBySpecimenDateRollingSum + dat$newCasesLFDOnlyBySpecimenDateRollingSum)/7
    ##dat$pctPCRpos = dat$PCRpos / dat$PCRtst
    if (bCorLFDDenom) dat$dT.NoRep = dat$dT.NoRep - dat$dC.LFD
    dat$pctPCRpos = (dat$newCasesPCROnlyBySpecimenDateRollingSum/7)/dat$dT.NoRep
    dat$pctPCRPpos = dat$uniqueCasePositivityBySpecimenDateRollingSum/100 -
        dat$newCasesLFDConfirmedPCRBySpecimenDateRollingSum/dat$uniquePeopleTestedBySpecimenDateRollingSum
    ##Note; pctPCRPposNEW is shown numberically to be equal to pctPCRpos
    dat$pctPCRPposNEW = dat$uniqueCasePositivityBySpecimenDateRollingSum/100 * 
         dat$newCasesPCROnlyBySpecimenDateRollingSum/( dat$newCasesPCROnlyBySpecimenDateRollingSum +  dat$newCasesLFDConfirmedPCRBySpecimenDateRollingSum)
    dat$pctPCRPunadj = dat$uniqueCasePositivityBySpecimenDateRollingSum/100
    ##dat$pctPCRpos = 0.01 * dat$uniqueCasePositivityBySpecimenDateRollingSum ##experimental
    dat = dat[!is.na(dat$pctPCRpos),]
    dat$t = as.numeric(dat$date - dat$date[1])    
    rownames(dat) = 1:dim(dat)[1]

    
    if (!bDebug) dat = dat[,c('date','t', 'pctPil1', 'pctPCRPunadj', 'pctPCRpos', 'pctPCRPpos')]
    
    return(dat)
}

###################################################################################################################
## return the England vaccination data from the file

getVaxEngland<-function(fname ='~/Downloads/englandVaccineAndMisc.csv')
{
    popEngland = 56.287e6 ## ONS most recent (24 June 2020) estimate for 2019.

    dat=read.csv(fname, stringsAsFactors=FALSE)

    dat$date = as.Date(dat$date, "%Y-%m-%d")
    if (any(colnames(dat) == "newCasesLFDOnlyBySpecimenDateRollingSum"))
    { ## get rid of unused miscillaneous non-vax fields
        dat = dat[,-which(colnames(dat) ==  "newCasesLFDOnlyBySpecimenDateRollingSum")] 
    }
    ## the following 4 additional data points (before the PHE time series starts) are from Our World in Data, UK.
    preDat = data.frame(date=as.Date(c('2020-12-13','2020-12-20','2020-12-27', '2021-01-03')), vax1=c(0.0013,0.0099, 0.0148, 0.0203), vax2=c(0,0,0,0)) 
    
    dat=dat[,-match(c('areaType','areaCode','areaName'), colnames(dat))]
    colnames(dat) = gsub(x=colnames(dat), pattern='cumPeopleVaccinatedFirstDoseByPublishDate', replacement='vax1')
    colnames(dat) = gsub(x=colnames(dat), pattern='cumPeopleVaccinatedSecondDoseByPublishDate', replacement='vax2')
    dat$vax1 = dat$vax1/popEngland
    dat$vax2 = dat$vax2/popEngland

    dat = dat[order(dat$date),]
    dat = dat[!is.na(dat$vax1) & !is.na(dat$vax2),] ##Empty cells mess up the below interpolation
    preDat = rbind(preDat, dat[1,])
  
    nPreDat2 = as.numeric(max(preDat$date) - min(preDat$date)) - 1
    newDates = as.Date(preDat$date[1] + (0:nPreDat2))
    preDat2 = data.frame(date=newDates,
                         vax1 = approx(x=preDat$date, y=preDat$vax1, xout=newDates)$y,  ##Interpolate the Our World In Data datapoints
                         vax2= approx(x=range(preDat$date), y=range(preDat$vax2), xout=newDates)$y ) ##extrapolate the first PHE data point 
    dat = rbind(preDat2, dat)    
    dat$vax12 = dat$vax2 + 0.8 * (dat$vax1 - dat$vax2) ##100% protection 2nd dose, 80% first dose
    rownames(dat) = 1:dim(dat)[1]
    return(dat)
}


#####################################################################################
## Run the analysis and draw the graph from the paper

getEnglandCorrected<-function
(
    startDate="2021-01-06",    	##Start Date: Corresponds to start date of 3rd England Lockdown
    startPeriod1="2021-03-08", 	##Default is when UK Gov reopened for in person learning
    dat=NULL,      		##This gives the option of passing a historic pre-computed data frame (table) to the analysis. 
    				##Note: dat uses an older data-collection scheme that would need extra work to document for you & can be provided on request
    nExtendPlot=10, 		##Number of additional points to extend (predict) the data
    bDoPlot=TRUE,   		##Optional to display the plot
    bRetData=TRUE,  		##Option to return the data matrix
    bOriginalModel = FALSE,	##Model from the 2021-04-07 paper, Depricated -- for comparison only
    bOriginalPCRestm = FALSE,	##Use original PCR estimate: only valid when providing input data matrix (see dat parameter)
    logGraph='y',   		##option to plot the graph on log y scale, leave empty, i.e. '' to not do this 
    nPoints=NA,     		##Default number of points to use in the plotting and regression, allows "back testing"
    useVax=12,      		## 1 = use only first vax, 2=use only second vax, 12=use both vaccinations in 80/20 ratio
    vaxDelay=7,    		## Mean Delay between vaccation administration & effective epidemiological immunity (differs from clinical certainty)
    bUsePCRP=FALSE, 		## Option to use PHEs PCR "People" (not tests) ratio with only a correction for LFD bias
    bUsePCRPunadj=FALSE,  	##Option to not adjust PHEs PCRP ratio for LFD bias, i.e. use as is
    bLogModel=TRUE, 		## For nls non-linear least squares models: T=minimize log of prediction error; F=min (non-log) prediction error
    bFindFPR=FALSE, 		## For nls models only, add in option to find the False Positive Ration
    bOnePeriodModel=FALSE,	## The default is a two period model now, but other uses will find one period better
    bTrace = FALSE, 		##Debugging switch for nls style models to trace (print) to possible non-convergence iterations
    bShowLinearModel=FALSE,	##For testing to prove lm (ordinary least squares regression) matches the nlm result 
    bCorLFDDenom=FALSE,		##Correct the PCR positivity denominator by removing LFD tests presumably submitted for PCR verification
    bPlotNewerData=TRUE,	##If the data is back tested (truncated), display newer data points for comparison
    bUseNewVaxData=FALSE,	##If false, extrapolate out the vax data, if true, use new vax data where available
    bPlotSpotImplied=FALSE	##The spot future spot rate is implied from the linear model for the mean vax, not used but may be interesting
)
{
    bNewModel = !bOriginalModel

    if (is.null(dat)) dat = getEnglandData(startDate=startDate, bCorLFDDenom=bCorLFDDenom)
    
    startPeriod1 = as.Date(startPeriod1)
    endPeriod0 = startPeriod1 - 1

    if (!any(colnames(dat) == 'pctPCRpos'))
    {
        if (bOriginalPCRestm)
        {
            dat$PCRtst = c(dat$dTsPCRma[-(1:2)], NA, NA)
            dat$PCRtst = dat$PCRtst - dat$dLFDPCRma - dat$dLFD2Xma
        }
        else
        {
            dat$dT.PCRP = dat$dTsPCRPma
            dat$dC.PCRP = dat$dCPCRma
            dat$dC.PCR = dat$dLFDPCRma + dat$dPCR2Xma
            dat$dT.NoRep = dat$dC.PCR/dat$dC.PCRP * dat$dT.PCRP
            dat$PCRtst = dat$dT.NoRep
        }
        dat$pctPil1 = dat$dTsPil1MA/dat$dTsPCRma
        dat$pctPil1 = c(dat$pctPil1[-(1:2)], NA, NA) 
        dat$pctPCRpos = dat$dPCR2Xma / dat$PCRtst
        dat$pctPCRPpos = dat$pctPCR7 - dat$dLFDPCRma/dat$dTsPCRma        
        dat$pctPCRPunadj = dat$pctPCR7
        dat = dat[!is.na(dat$pctPCRpos),]
        dat = dat[dat$date>=as.Date(startDate),]
        dat$t = as.numeric(dat$date - dat$date[1])    
        rownames(dat) = 1:dim(dat)[1]
        dat = dat[,c('date','t', 'pctPil1', 'pctPCRpos', 'pctPCRPpos', 'pctPCRPunadj')]        
    }

    datAll = dat ##datAll is all the data without trunctation so can compare how predictions are doing to actual data
    
    if (bUsePCRP) dat$pctPCRpos = dat$pctPCRPpos
    if (bUsePCRPunadj) dat$pctPCRpos = dat$pctPCRPunadj
    if (is.null(nPoints)) nPoints = NA
    if (!is.na(nPoints)) dat = dat[1:(min(dim(dat)[1], nPoints)),]  ##Truncate the data to nPoints
    
    datVax = getVaxEngland()
    
    datVax$vax = datVax$vax12
    if (useVax == 1) datVax$vax = datVax$vax1
    if (useVax == 2) datVax$vax = datVax$vax2
    datVax$vax[(1+vaxDelay):dim(datVax)[1]] = datVax$vax[1:(dim(datVax)[1]-vaxDelay)]
    dat$vax = rep(0, dim(dat)[1]) ## Empty place to put vax data
    datAll$vax = rep(0, dim(datAll)[1])
    
    ##Read the vax data into dat
    for (i in 1:dim(datVax)[1])
    {
        bCondition=any(datVax$date[i] == dat$date, na.rm=T)
        if (bCondition) dat$vax[dat$date == datVax$date[i]] = datVax$vax[i]
        bCondition=any(datVax$date[i] == datAll$date, na.rm=T)
        if (bCondition) datAll$vax[datAll$date == datVax$date[i]] = datVax$vax[i]
    }
    dat$vaxSpot = dat$vax ###For comparison, but not used
    datAll$vaxSpot = datAll$vax
   
    for (i in dim(dat)[1]:1) dat$vaxArith[i] = mean(dat$vaxSpot[1:i])  ##The effective integrated vax is the mean over period
    for (i in dim(dat)[1]:1) dat$vaxGeom[i] = exp(mean(log(dat$vaxSpot[1:i])))

    for (i in dim(datAll)[1]:1) datAll$vaxArith[i] = mean(datAll$vaxSpot[1:i])  ##The effective integrated vax is the mean over period
    for (i in dim(datAll)[1]:1) datAll$vaxGeom[i] = exp(mean(log(datAll$vaxSpot[1:i])))
    
    dat$vax = dat$vaxArith
    datAll$vax = datAll$vaxArith

    
    
    start = c(I0 = dat$pctPCRpos[1], r0 = 0.01, FPR = 0.0002, kvax=0.01)
    lower = c(I0 = 1e-8, r0 = -0.1, FPR = 0.0000, kvax=0)
    upper = c(I0 = 1, r0 =  0.5, FPR = 0.1, kvax=5)
        
    t1 = as.numeric(endPeriod0 - dat$date[1])    
    
    if (bNewModel)
        {
            iFPR = which(names(start)=='FPR')
            start = start[-iFPR]
            lower = lower[-iFPR]
            upper = upper[-iFPR]
            start = c(start, r1 = 0.01)
            lower = c(lower, r1 = -0.5)
            upper = c(upper, r1 = 0.5)
            start = start[order(names(start))]
            lower = lower[order(names(lower))]                         
            upper = upper[order(names(upper))]
        }
        
    ##LOG LINEAR VERSION
    ## Note: CANNOT linearized FPR due to sum of the two halfs! i.e. log(exp(A) + B) cannot simplify
    modelLinear = NULL
    if (bShowLinearModel && bLogModel & bNewModel) ##THIS WORKS I0 = exp(regression constant), 
        modelLinear = formula(log(pctPCRpos) ~
                                  I(-(vax)*t) +  ##kvax will be this coefficient
                                  I(-pmin(t, t1)) + ##r0 will be this coefficient 
                                  I(-pmax(0, t - t1)))  ##r1 will be this coefficient
 
    if (!bFindFPR && bLogModel && bNewModel)
        modelFormula = formula(log(pctPCRpos) ~ log(I0 *  exp( -(r0  * pmin(t, t1))  -(r1 * pmax(0, t-t1)) - kvax*vax*t)) )
   
    if (bFindFPR && bLogModel && bNewModel)
        modelFormula = formula(log(pctPCRpos) ~ log((1-FPR) * (I0 *  exp( -(r0  * pmin(t, t1))  -(r1 * pmax(0, t-t1)) - kvax*vax*t)) + FPR ) )

    if (!bFindFPR && !bLogModel && bNewModel)
        modelFormula = formula(pctPCRpos ~ I0 *  exp( -(r0  * pmin(t, t1))  -(r1 * pmax(0, t-t1)) - kvax*vax*t) )
   
    if (bFindFPR && !bLogModel && bNewModel)
        modelFormula = formula(pctPCRpos ~ (1-FPR) * (I0 *  exp( -(r0  * pmin(t, t1))  -(r1 * pmax(0, t-t1)) - kvax*vax*t)) + FPR )
    
    

    if (bFindFPR && bNewModel)
    {
        start = c(start, FPR = 0.001)
        lower = c(lower, FPR = 0.000)
        upper = c(upper, FPR = 0.10)
    }
    

    if (bOnePeriodModel)
      {
        ir1 = which(names(start)=='r1')
        if (any(ir1))
            {
                start = start[-ir1]
                lower = lower[-ir1]
                upper = upper[-ir1]
            }
      }
      

    if (bOnePeriodModel & bFindFPR & bLogModel)
        modelFormula = formula(log(pctPCRpos) ~ log((1-FPR) * (I0 *  exp( -(r0 + kvax*vax)*t)) + FPR ) )
                               
    if (bOnePeriodModel & !bFindFPR & bLogModel)
        modelFormula = formula(log(pctPCRpos) ~ log(I0 *  exp( -(r0 + kvax*vax)*t)) )
                               
    if (bOnePeriodModel & bFindFPR & !bLogModel)
        modelFormula = formula(pctPCRpos ~ (1-FPR) * (I0 *  exp( -(r0 + kvax*vax)*t)) + FPR  )
                               
    if (bOnePeriodModel & !bFindFPR & !bLogModel)
        modelFormula = formula(pctPCRpos ~ I0 *  exp( -(r0 + kvax*vax)*t) )

    if (bOnePeriodModel & bShowLinearModel)
    {
        modelLinear = formula(log(pctPCRpos) ~
                                  I(-(vax)*t) +  ##kvax will be this coefficient
                                  I(-t) ) ##r0 will be this coefficient 
 
    }

    if (bOriginalModel)
    { ##Model from original paper -- OVERRIDES ANY OTHER MODEL
        modelFormula = formula(log(pctPCRpos) ~ log((1 - FPR) * I0 * exp( - (r0 + kvax*(vax-vax0)* t) * t) + FPR))
        bLogModel = TRUE
    }

    
    iFPR = which(names(start) == 'FPR')

    if (useVax == 0)
    {
        kvax = 0
        ikvax = which(names(start) == "kvax")
        start = start[-ikvax]
        lower = lower[-ikvax]
        upper = upper[-ikvax]
    }
    
    vax0 = dat$vax[1]
    vax1 = dat$vax[dat$date == endPeriod0]
    vaxSpot1 = dat$vaxSpot[dat$date == endPeriod0]
    vaxSpot0 = dat$vaxSpot[1]
    model = nlsLM(formula = modelFormula ,
                    start=start,
                    lower=lower,
                    upper=upper,
                    algorithm='port',
                    trace = bTrace,
                    control = nls.control(maxiter=999),
                    na.action = na.omit,
                    data=dat)

    print(summary(model), signif.stars = FALSE)
    printf("-------------------------------\n")
    printf("NLS Model Upper/Lower Bounds:\n")
    print(format(scientific=FALSE, data.frame(rbind(upper,lower))))
    printf("-------------------------------\n")

    if (!is.null(modelLinear))
        print(summary(lm(modelLinear, data=dat)))

    tMax = max(dat$t)
    tMax0 = dat$t[dat$date == endPeriod0]
    tMax1 = tMax - tMax0
    vaxFinal = dat$vax[dim(dat)[1]]
    vaxFinal0 = dat$vax[dat$date == endPeriod0]
    vaxFinal1 = vaxFinal
    vaxSpotFinal = dat$vaxSpot[dim(dat)[1]]
    vaxSpotFinal0 = dat$vaxSpot[dat$date == endPeriod0]
    vaxSpotFinal1 = vaxSpotFinal
    vaxMeanPrior = dat$vax[dim(dat)[1]-1]
    vaxMeanPrior1 = vaxMeanPrior
    vaxMeanPrior0 = dat$vax[which(dat$date == endPeriod0)]

    mod = as.list(model$m$getPars())
    mod$rEnd = mod$r0 + 2 * mod$kvax * (vaxFinal - vax0)*tMax + mod$kvax * tMax^2/(tMax-1) * (vaxSpotFinal - vaxMeanPrior)    
    if (bNewModel)
    {
        mod$rEnd = NA
        mod$rEnd0 = mod$r0 + mod$kvax * vaxFinal0 + mod$kvax * tMax0 * (vaxSpotFinal0 - vaxMeanPrior0)/(tMax0 - 1)
        mod$rEnd1 = mod$r1 + mod$kvax * vaxFinal1 + mod$kvax * tMax  * (vaxSpotFinal1 - vaxMeanPrior1)/(tMax  - 1)
    }
    modCoefs = summary(model)$coefficients
    modSigma = as.list(modCoefs[,'Std. Error'])
    if (useVax == 0) modSigma$kvax = NA
    
    if (!bNewModel)      
        modSigma$rEnd = sqrt(modSigma$r0^2 + (2 *tMax * (vaxFinal - vax0) * modSigma$kvax)^2
                             + (modSigma$kvax * tMax^2/(tMax-1) * (vaxFinal - vaxMeanPrior))^2 )
    if (bNewModel)      
        modSigma$rEnd0 = sqrt(modSigma$r0^2 + ((vaxFinal0 - vax0) * modSigma$kvax)^2
                             + (modSigma$kvax * tMax0 * (vaxFinal0 - vaxMeanPrior0))^2 )
    if (bNewModel)      
        modSigma$rEnd1 = sqrt(modSigma$r1^2 + ((vaxFinal1 - vax1) * modSigma$kvax)^2
                             + (modSigma$kvax * tMax1 * (vaxFinal1 - vaxMeanPrior))^2 )

    mp = lapply(mod, function(x) {100*x}) ##For percent printout
    ms = lapply(modSigma, function(x) {100*x})
    period=range(dat$date)
    nEnd = dim(dat)[1] ##remember this for later

    printf("Start Period  %s to %s End Period  : %3d days\n", period[1], period[2], tMax + 1)
    if (bNewModel) printf("Start Period0 %s to %s End Period0 : %3d days\n",
                          period[1], endPeriod0, as.numeric(endPeriod0 - period[1]) + 1)
    if (bNewModel) printf("Start Period1 %s to %s End Period1 : %3d days\n",
                          startPeriod1, period[2], as.numeric(period[2] - startPeriod1) + 1)
    printf("Mean Vaccination: Start:   %s %6.3f%%\n", period[1], 100*vax0)
    printf("Mean Vaccination: Start to %s %6.3f%%\n", endPeriod0, 100*vaxFinal0)
    printf("Mean Vaccination: Start to %s %6.3f%%\n", period[2], 100*vaxFinal1)
    printf("-------------------------------------------------------------\n")
    printf("Initial Prevalence            I0 = %7.3f%% (CI %7.3f - %7.3f%%)\n", mp$I0, mp$I0-2*ms$I0, mp$I0+2*ms$I0)
    printf("Decay per Mean Vaccinated   kvax = %7.3f%% (CI %7.3f - %7.3f%%)\n", mp$kvax, mp$kvax-2*ms$kvax, mp$kvax+2*ms$kvax)
    printf("Initial Daily Decay Rate      r0 = %7.3f%% (CI %7.3f - %7.3f%%)\n", mp$r0, mp$r0-2*ms$r0, mp$r0+2*ms$r0)
    if (bNewModel)
        printf("Initial Daily Decay Rate      r1 = %7.3f%% (CI %7.3f - %7.3f%%)\n", mp$r1, mp$r1-2*ms$r1, mp$r1+2*ms$r1)
    if (!bNewModel)
        printf("End Period Decay Rate       rEnd = %7.3f%% (CI %7.3f - %7.3f%%)\n", mp$rEnd, mp$rEnd-2*ms$rEnd, mp$rEnd+2*ms$rEnd)
    if (bNewModel)
        {
            printf("End Period0 Decay Rate     rEnd0 = %7.3f%% (CI %7.3f - %7.3f%%)\n", mp$rEnd0, mp$rEnd0-2*ms$rEnd0, mp$rEnd0+2*ms$rEnd0)
            printf("End Period1 Decay Rate     rEnd1 = %7.3f%% (CI %7.3f - %7.3f%%)\n", mp$rEnd1, mp$rEnd1-2*ms$rEnd1, mp$rEnd1+2*ms$rEnd1)
        }
    printf("False Positive Rate          FPR = %7.3f%% (CI %7.3f - %7.3f%%)\n", mp$FPR, mp$FPR-2*ms$FPR, mp$FPR+2*ms$FPR)

    nPoints = dim(dat)[1]
    df = nPoints - 3 - 1
    if (bLogModel)
    {
        unexplainedPCR = var(predict(model) - log(dat$pctPCRpos))*(nPoints - 1)/df    ##Correct R's variance for degrees of freedom
        varPCR = var(log(dat$pctPCRpos))
    }
    else
    {
        unexplainedPCR = var(predict(model) - (dat$pctPCRpos))*(nPoints - 1)/df    ##Correct R's variance for degrees of freedom
        varPCR = var((dat$pctPCRpos))
    }
    rsquaredPCR = (varPCR-unexplainedPCR)/varPCR
    rsquaredPCRadj = 1 - (1 - rsquaredPCR) * (nPoints - 1)/(df)

    ##Note: this is a worst case to compute final prevalence CI based on a 2 std error in the prediction
    ##Applied on a 2 standard error FPR, i.e. correlated in the worst way.
    if (bLogModel) 
        fExpOrNot = exp
    else
        fExpOrNot = function(x) {x}
    Iend = (fExpOrNot(predict(model))[nPoints] - mod$FPR)/(1 - mod$FPR)
    IendMax = (Iend*fExpOrNot(2*sqrt(unexplainedPCR)) + (2*modSigma$FPR))/(1+2*modSigma$FPR)
    IendMin = (Iend*fExpOrNot(-2*sqrt(unexplainedPCR)) - (2*modSigma$FPR))/(1-2*modSigma$FPR)
    IendMin = max(0, IendMin)
      
    printf("Final Prevelence            Iend = %7.3f%% (CI %7.3f - %7.3f%%)\n", 100*Iend, 100*IendMin, 100*IendMax)
    printf("PCR Fit Statistics var = %7.5f SD resid = %7.5f rSquared = %7.5f rSquared Adj = %7.5f\n",
           varPCR, sqrt(unexplainedPCR), rsquaredPCR, rsquaredPCRadj)

        
    if (nExtendPlot>0)
    {
        nDat = dim(dat)[1]
        iExtend = 1:nExtendPlot
        dateMax = max(dat$date)
        tMax = max(dat$t)

        vaxModelDays = max(0,(nDat - 14)):nDat
        ##linearly extrapolated vaccines based on linear regression of last 14 days...
        if (bOriginalModel) vaxModelDays = max(0,(nDat - 30)):nDat
        vaxModel = lm(vax~date, data=dat[vaxModelDays,])
        vaxSpotModel = lm(vaxSpot~date, data=dat[vaxModelDays,])
        vaxGeomModel = lm(vaxGeom~date, data=dat[vaxModelDays,])
        vaxArithModel = lm(vaxArith~date, data=dat[vaxModelDays,])


        datesExtended = as.Date(iExtend+dateMax)
        dataNA = rep(NA, nExtendPlot)
        extendedDataNA = data.frame(date=datesExtended, matrix(NA, nExtendPlot, dim(dat)[2]-1))
        vaxPredict = predict(vaxModel, newdata=extendedDataNA)
        nPredictDays = length(predict(vaxModel))
        ## then use the new vax data to make the predictions so can check how model performs with actual vax data
        ## Note: there will be a small non-linearity blip as the model transitions from the last vax datapoint to the 1st linear regression prediction
        ## This has no effect on the current modelled data stream.
        if (bUseNewVaxData)
        {
            vaxPredict = dataNA
            for (i in iExtend)
            {
                iDate = which(datAll$date == as.Date(i+dateMax))
                if (any(iDate)) vaxPredict[i] = datAll$vax[iDate] 
            }
        }
       
        dat =  rbind(dat, data.frame(date=as.Date(iExtend+dateMax),
                                     t=tMax+iExtend,
                                     pctPil1=rep(dat$pctPil1[nDat], nExtendPlot),
                                     pctPCRpos=dataNA,
                                     pctPCRPpos = dataNA,
                                     pctPCRPunadj = dataNA,
                                     vax=vaxPredict,
                                     vaxSpot= dataNA,
                                     vaxArith= predict(vaxArithModel, newdata=extendedDataNA),
                                     vaxGeom=predict(vaxGeomModel, newdata=extendedDataNA)                                     
                                     )
                     )
        rownames(dat) = 1:dim(dat)[1]
    }

    ##Make vaxSpot predictions (for plotting) consistent with with vax predictions
    for (i in sort(which(is.na(dat$vaxSpot) & !is.na(dat$vax)))) 
        {
            dat$vaxSpot[i] = dat$vax[i] * i - dat$vax[i-1] * (i - 1)            
        }
 

    dat$fit = fExpOrNot(predict(model, newdata=dat))
    if (any(names(mod) == 'FPR'))
        dat$FPR = mod$FPR
    else
        dat$FPR = 0
    
    
    dat$I = (dat$fit - dat$FPR)/(1 - dat$FPR)
    
     
    ##yrange = range(na.rm=TRUE, c(dat$pctPCRpos, dat$fit, dat$I))
    ##These ranges are set manually by trial and error to make the graph use all the space in the paper
    yrange = c(0, 0.20)
    if (bOriginalModel) yrange = c(0, 0.16)
    ylabs = c(0.02 * (0:10))

    if (logGraph == 'y')
        {
            yrange[1] = 0.004
            ylabs = c(0.004, 0.006, 0.008, 0.010, ylabs)

            if (min(dat$I, na.rm=T)<0.004)
            {
                yrange[1] = 0.001
                ylabs = c(0.001, 0.002, ylabs)
            }
        }
            
    vaxMultiplier = 3
    if (bOriginalModel) vaxMultiplier = 3
    vaxRange = vaxMultiplier*yrange
    
    szSubtitle = sprintf("%s%s%s%s",
                         "X's: PCR Positivity, ",
                         "Black Line: Fitted Positivity, ",
                         "Black Dashes: Fitted FPR, ",
                         "Red: Fitted Prevalence"
                         )
    szSubtitle = ""                   
    
    if (bDoPlot)
    {
        par(mar=c(5, 4, 4, 5)+0.1)
        with(dat, plot(pctPCRpos~date, ylim=yrange, log=logGraph, type='p', pch='x', col='black',
                       yaxt='n', ylab="", xlab="",
                       main="England PCR Positivity Prevalence & Vaccination Immunity",
                       sub=szSubtitle)
             )

        axis(2, at=ylabs, cex.axis=0.88, las=1, mgp=c(3.5,0.7,0),labels=sprintf("%.1f%%", 100*ylabs))
        axis(4, at=ylabs, cex.axis=0.88,las=1, mgp=c(3.5,0.7,0),labels=sprintf("%.1f%%", 100*ylabs*vaxRange[2]/yrange[2]))
        mtext(side=1, line=2.25, "Date (2021)")
        mtext(side=2, line=2.7, "Positivity Data (X's=Old O's=New), Model Fit (Lines:  Red=Old Blue=New)")
        mtext(side=4, line=2.9, "Mean Effective Vaccine Immunity (Green=Original Data & Estimate, Blue=New Data)")
        lines(dat$fit~dat$date)
        lines(dat$I~dat$date, col='darkred')
        if (bPlotSpotImplied) lines((yrange[2]/vaxRange[2]*dat$vaxSpot)~dat$date, col='darkgreen')
        lines((yrange[2]/vaxRange[2]*dat$vax)~dat$date, col='darkgreen')
        abline(h=mod$FPR, lty="longdash", col='black')
        abline(h=ylabs, col='black', lty="dotted")
        dateHashes = as.Date(sprintf("2021-%02d-01", c(1:12)))
        abline(v=dateHashes, col='black', lty="dotted")
        text(cex=0.9, x=min(dat$date)+1, y=mod$FPR+0.001, adj=0, "False Positive Rate") ##Labe
        if (bNewModel) abline(v=endPeriod0, col=rgb(0.2,0.2,0.2),lty="dashed")
        if (bPlotNewerData)
        {
            with(datAll[-(1:nPoints),], points(pctPCRpos~date, col='darkblue'))
            if(bPlotSpotImplied) with(datAll[-(1:nPoints),], points(vaxSpot/vaxMultiplier~date, type='l', col='darkblue'))
            with(datAll[-(1:nPoints),], points(vax/vaxMultiplier~date, type='l', col='darkblue', lwd=2))
        }
    }

    ##Export to dat the newer vaxccine data for comparison on back tested model run
    dat$vaxArithNew = dat$vaxSpotNew = rep(NA, dim(dat)[1])
    nSmaller = min(dim(dat)[1], dim(datAll)[1])
    dat$vaxArithNew[1:nSmaller]= datAll$vax[1:nSmaller]
    dat$vaxSpotNew[1:nSmaller] = datAll$vaxSpot[1:nSmaller]
    
    if (bRetData) return(dat)
    
}
```
